# Estradiol’s Effect on Reward-Related Functional Connectivity in Perimenopause-Onset Major Depressive Disorder

**DOI:** 10.1101/2025.07.29.25332383

**Authors:** Megan Hynd, Kathryn Gibson, Melissa Walsh, Julianna Prim, Tory Eisenlohr-Moul, Erin Walsh, Lauren Schiff, Gabriel Dichter, Crystal Schiller

## Abstract

**Background:** Hormone replacement therapy with estradiol (E2), the most common form of estrogen, is an effective intervention for affective symptoms associated with perimenopause-onset major depressive disorder (PO-MDD). However, the precise neural mechanisms by which E2 influences these symptoms remain unclear. This study aimed to investigate changes in reward-related frontostriatal connectivity following E2 administration for PO-MDD using fMRI with a monetary incentive delay task.

**Methods:** Fifteen participants with PO-MDD and 16 control participants underwent three weeks of transdermal E2 administration to evaluate its impact on mood symptoms and frontostriatal connectivity. Functional MRI scans were conducted before and after the administration of E2. Changes in functional brain connectivity related to reward anticipation were examined using the CONN toolbox. Depression symptoms were assessed with the IDAS Dysphoria and the MASQ-AD scales.

**Results:** The PO-MDD group exhibited altered connectivity compared with controls during reward anticipation prior to E2 administration. During reward anticipation, they exhibited decreased connectivity from striatal and cortical seed regions, including the caudate, putamen, nucleus accumbens, and insula seeds. E2 administration increased connectivity during reward anticipation across many of these seed regions in the PO-MDD group but not in the control group. Within the PO-MDD group, changes in functional connectivity following E2 administration predicted reduced dysphoria, with increased connectivity between the insula– angular gyrus linked to greater symptom reduction over three weeks. In contrast, decreased insula–inferior temporal gyrus connectivity was associated with greater symptom reduction over three weeks.

**Conclusion:** The PO-MDD group exhibited reduced functional connectivity during reward anticipation compared with controls, with distinct patterns of connectivity with striatal and limbic seeds, including the caudate nucleus, putamen, nucleus accumbens, and insula. E2 administration generally increased connectivity between striatal and limbic seed regions and the cortex. Additionally, changes in connectivity during reward anticipation predicted greater reductions in dysphoria following E2 administration. These findings highlight the potential significance of reward-related functional connectivity for explaining the triggering of PO-MDD and potential mechanisms underlying the antidepressant effects of E2.

## INTRODUCTION

Perimenopause marks the transition to menopause and is linked to a greater risk of anhedonia and major depressive disorder (MDD)(Dibonaventura et al., 2012; Wariso et al., 2017). Fluctuations in hormone levels, especially estradiol (**E2**), during this time are thought to contribute to the onset of major depressive disorder related to perimenopause (**PO-MDD**)(Bromberger & Epperson, 2018; Deecher et al., 2008; Gordon et al., 2016, 2016). During perimenopause, women experience more pronounced daily changes in E2—both higher peaks and lower troughs—compared with a typical menstrual cycle. This unstable fluctuation in E2 significantly impacts brain neurochemistry, structure, and function, potentially disrupting reward processing and resulting in anhedonia, which is a key symptom of MDD (Gordon & Sander, 2021).

Anhedonia describes a diminished capacity to experience pleasure or show interest in previously enjoyable activities and is experienced by up to 70% of individuals with MDD (Cao et al., 2019). Central to this symptom is the dysregulation of the brain’s reward processing systems. Individuals with MDD often exhibit altered activity in key reward-related brain regions, such as the striatum, anterior cingulate cortex (ACC), and the prefrontal cortex (Nagy et al., 2020; Whitton et al., 2023). These areas are crucial for evaluating reward values, anticipating future rewards, and motivating behaviors to obtain rewards. Hypoactivity to reward cues has generally been observed in the striatum, ACC, and orbitofrontal cortex, while the medial frontal pole, ventromedial prefrontal cortex, and dorsolateral prefrontal cortex appear to be hyperactive as a potential compensatory mechanism (Pizzagalli, 2022). The striatum specifically plays a pivotal role in reward anticipation, with individuals with MDD showing diminished striatal response during reward anticipation (Whitton et al., 2015). Further, aberrant frontostriatal functional connectivity characterizes anhedonia and may represent a marker of vulnerability to later MDD (Pan et al., 2022).

The activity and connectivity of brain reward circuitry are highly susceptible to fluctuations in E2 (Andreano & Cahill, 2009; Dreher et al., 2007; Kenna et al., 2009). These regions express high concentrations of estrogen receptors (Brinton et al., 2015), supporting a mechanistic role for E2 in modulating reward-related processing. Indeed, increased endogenous E2 across the menstrual cycle is associated with increased putamen-thalamic connectivity (Hidalgo-Lopez et al., 2020). In a single densely-sampled subject, endogenous E2 fluctuations across the cycle changed the functional architecture of intrinsic brain networks, including between reward-related circuitry nodes (Pritschet et al., 2020). Further, exogenous administration of E2 produces increased functional connectivity between the basal ganglia and the thalamus (Kenna et al., 2009).

The robust influence of E2 on reward circuitry, combined with the pronounced fluctuations in E2 during the transition to menopause beyond what occurs in normal menstrual cycles, provides strong evidence that perimenopausal disruptions in E2 may trigger anhedonia and the development of MDD by disrupting reward-related functional connectivity. Short-term transdermal E2 administration is thought to alleviate perimenopause-onset MDD symptoms (Schiller et al., 2016) by changing activation of key nodes in the reward system (e.g., caudate nucleus, putamen, and nucleus accumbens). However, no studies to date have investigated reward-related functional connectivity changes that may underlie the clinical efficacy of transdermal E2 in PO-MDD.

To address this gap in the literature, we examined changes in task-based functional connectivity during a three-week trial of E2 administration in women with and without perimenopause-onset MDD (PO-MDD). Participants in this trial were healthy, unmedicated, perimenopausal women ages 44-55 with (n=15) or without (n=16) current MDD. For the PO-MDD group, the onset of MDD coincided with the transition to menopause, and there were no other psychiatric illnesses within the two years prior to study participation. The control group had no history of psychiatric illness. Both groups received three weeks of transdermal E2 (see *Hormone Administration and Monitoring* for details).

The current secondary analysis of existing data (E. C. Walsh et al., 2025) had three specific aims: The first aim was to examine baseline (pre-E2) group differences in functional connectivity during reward anticipation (between the PO-MDD and control groups). Based on previous findings of decreased neural response to reward in individuals with MDD (Bore et al., 2023), it was hypothesized that the PO-MDD group would display hypoconnectivity in reward-related circuitry during reward anticipation compared with the control group at baseline (pre-E2). The second aim was to test for group differences in pre-post E2 changes in functional connectivity during reward anticipation. It was hypothesized that the PO-MDD patients would show greater pre-post E2 increases in connectivity during reward anticipation between the striatum, insula, and other regions implicated in reward processing impairments in MDD. This hypothesis was predicated on the central role of the striatum in reward processing and the modulatory effects of estrogen on striatal dopamine transmission (Chavez et al., 2010). The third aim examined how changes in functional connectivity during reward anticipation related to symptom reduction in the PO-MDD group. It was hypothesized that changes in task-related functional connectivity during reward anticipation would moderate the relationship between E2 administration and the trajectory of depressive symptom reduction in the PO-MDD group.

## METHODS

### Participants

Forty-three perimenopausal women were recruited through social media ads, community flyers, and mass emails. The participants were divided into two groups: (1) those with PO-MDD (n = 20) and (2) those who had never experienced Axis I disorders (control group, n = 23). As described elsewhere (E. C. Walsh et al., 2025), inclusion criteria for both groups required participants to be 44-55 years old and in the late stage of perimenopause, defined by either ≥ 60 days of amenorrhea or two missed menstrual cycles per the Stages of Reproductive Aging Workshop (Stage −1) (Siobán D. Harlow et al., 2012). The exclusion criteria included current medication use, BMI less than 18 or greater than 30 kg/m2, pregnancy or breastfeeding, hormonal contraindications (such as cardiovascular disease or a personal or family history of breast cancer), MRI contraindications, recurrent major depressive MDD, or MDD unrelated to perimenopause. The PO-MDD group consisted of 16 participants with usable fMRI data at both time points, while the control group had 18 participants with usable data at both assessments. Usable data were defined as falling within the interquartile range on quality assurance measures, including the proportion of valid scans, mean motion, and global signal change. Data outside the first and third quartiles were excluded.

### Procedures

#### Hormone Administration and Monitoring

All participants were administered transdermal E2 (100 µg/day via Climara® patch) (Bayer HealthCare Pharmaceuticals, 2007) for three weeks after their baseline (“pre-E2”) MRI session. Following the post-E2 MRI session, they received an additional week of combined transdermal E2 (100 µg/day via Climara® patch) and micronized progesterone (200 mg/day via Prometrium®) to induce menstruation and minimize the risk of endometrial hyperplasia. Hormone adherence was verified through plasma E2 and progesterone levels assessed during each imaging session and at weekly clinic visits (see Figure 1).

**Figure 1.**
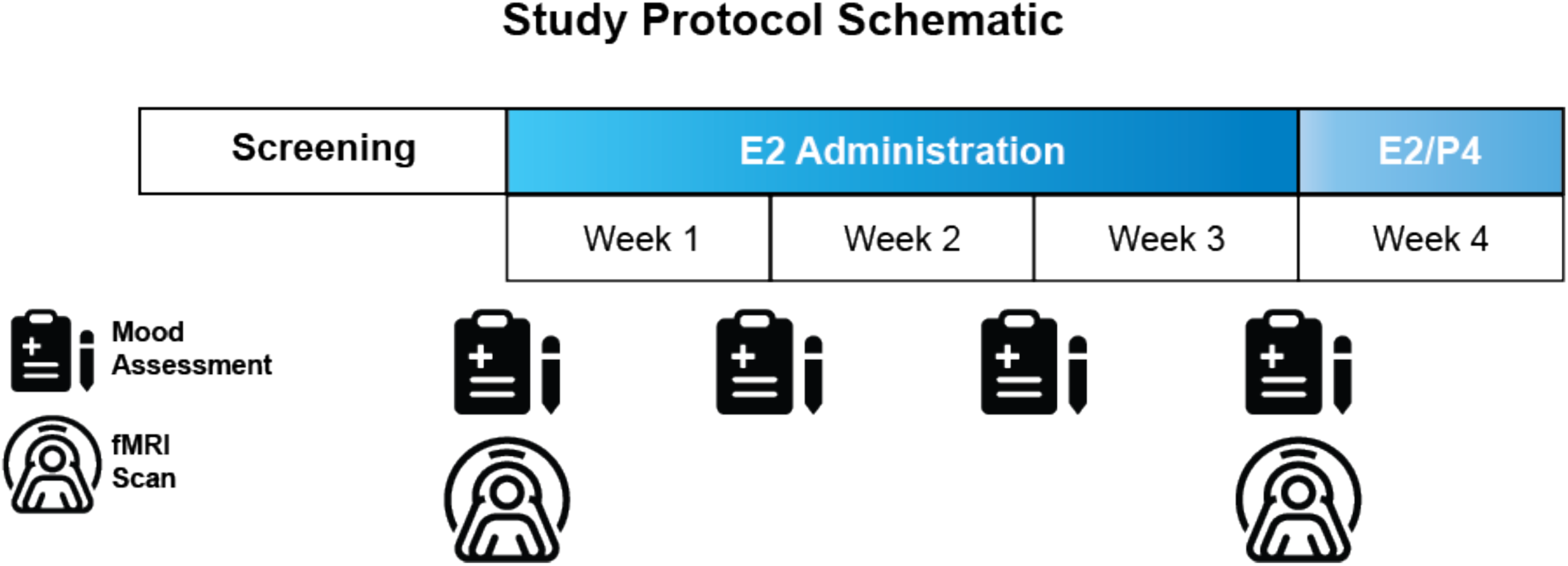
E2 administration protocol. Imaging was acquired at pre-E2 (intervention week 0) and post-E2 follow-up (intervention week 3). E2: estradiol. P4: progesterone.

**Figure 2.**
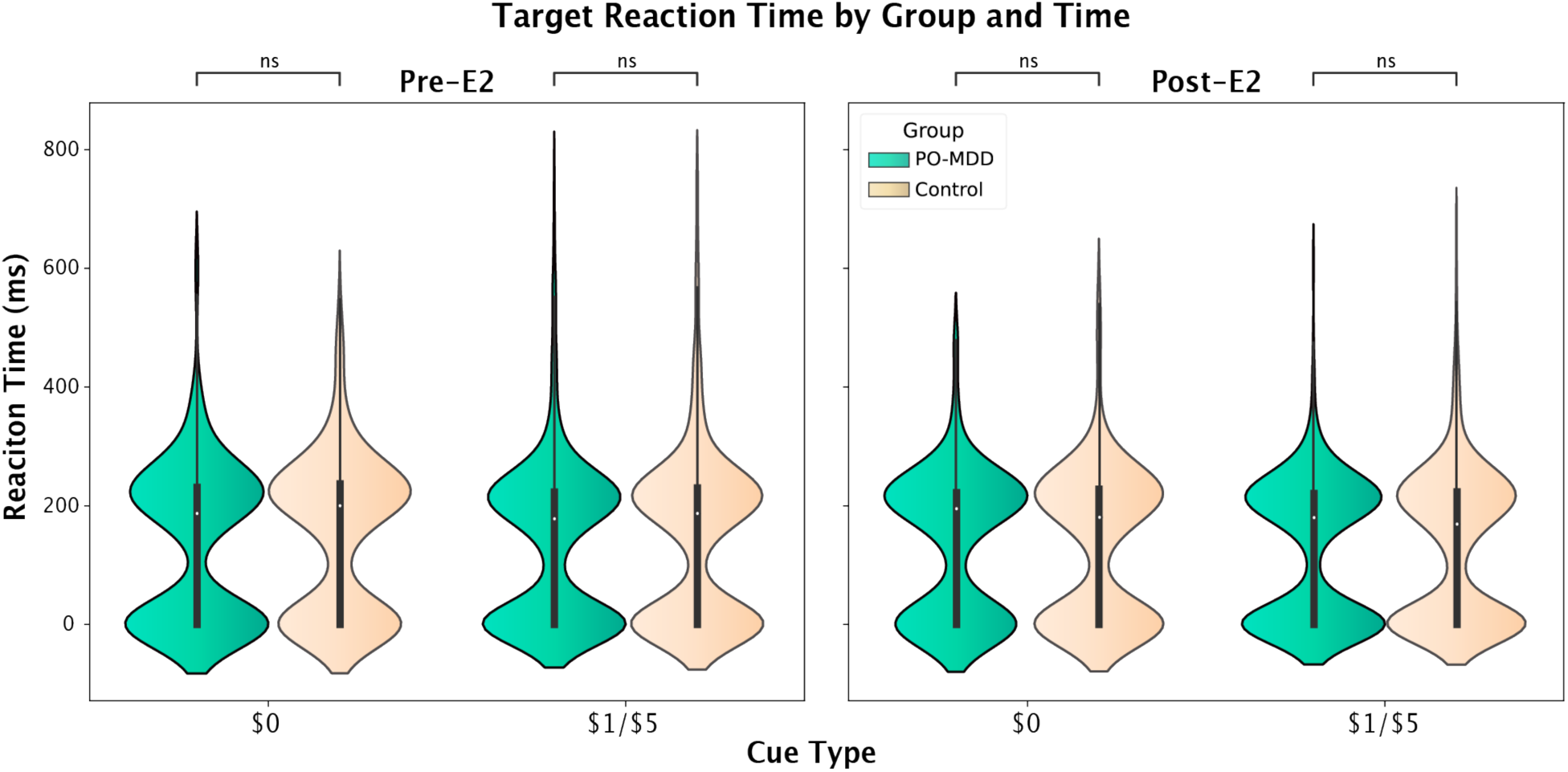
Violin plots for pre-E2 and post-E2 target reaction times separated by group and cue type. ns = non-significant.

#### Clinical Assessments

Mood symptoms were evaluated at the initial eligibility assessment, prior to the pre-E2 and post-E2 MRI, and during weekly clinic visits. The Structured Clinical Interview for DSM-IV-TR for Axis I Disorders (SCID-IV) was used to assess psychiatric illness during the initial eligibility assessment (First & Gibbon, 2004).

The following measures were administered at the pre-E2 eligibility assessment, during subsequent imaging sessions, and weekly clinic visits: 1) The Mood and Anxiety Symptom Questionnaire – Anhedonic Depression Subscale (**MASQ-AD**) consists of 22 items designed to evaluate anhedonia (Watson & Clark, 2012); prior studies on patients with MDD have linked MASQ-AD scores to reduced striatal response to reward (Pizzagalli et al., 2009). 2) The Inventory of Depression and Anxiety Symptoms (**IDAS**) is a 64-item scale that evaluates anxiety and depression through 10 subscales, including in reproductive-related mood disorders (Watson et al., 2007).

#### Imaging Acquisition

Scans were acquired at the University of North Carolina at Chapel Hill using a 3T TIM Trio Scanner. T1-weighted images were obtained using an MPRAGE sequence with these parameters: echo time (TE) = 3.16ms, repetition time (TR) = 2400ms, FOV = 256mm, acquisition matrix = 256 x 224 x 160mm, flip angle = 8°, voxel size = 1 x 1 x 1mm, 160 axial slices. Functional blood-oxygen-level-dependent (BOLD) images were acquired using a multi-slice, interleaved pulse sequence with the following parameters: TE = 25ms, TR = 2000ms, FOV = 256mm, acquisition matrix = 256 x 232 x 144mm, flip angle = 80°, voxel size = 4 x 4 x 4 mm; 36 axial slices.

The Monetary Incentive Delay (MID) task was used to probe reward circuitry function (Knutson et al., 2001). Stimuli were presented using E-Prime (Psychology Software Tools Inc. Pittsburgh, PA) on a mirrored screen. Participants received training outside the scanner to ensure understanding of the cues and were informed they would earn cash based on their performance before entering the scanner. They then completed one practice session outside the scanner, followed by two runs in the scanner, with 144 trials total. In each trial, one of seven cue shapes was displayed to signal either that no response was necessary or to indicate outcomes of low, medium, or high gain or loss (cue; 2000 ms). This was followed by a crosshair displayed for a variable interval (anticipation; 2000-2500 ms). Finally, a target was shown for a variable duration, prompting participants to press a button as quickly as possible (target; up to 500 ms) (Knutson et al., 2001). After the target presentation, feedback was provided showing the money earned or lost in that trial, along with the overall amount gained throughout the run (outcome; 3000 ms). The task was designed to ensure timely responses to targets in two-thirds of the trials, irrespective of reaction times. fMRI volume acquisitions were time-locked to cue offsets and outcome displays. (Knutson et al., 2001).

As described previously (Knutson et al., 2008), cues identified potential gain (28 trials, indicated by circles), potential loss trials (26 trials, indicated by squares), or no response required (18 trials, indicated by triangles). Each 9-minute run included ten $5 (“high”), nine $1 (“low”), and nine $0 (“non-win”) Gain trials. There were also Loss trials that included-$5 (“high”),-$1 (“low”), and $0 (no loss) Loss trials. The number of lines within each shape denoted the magnitude of the gain or loss: $0 (no lines), $1 (one line), or $5 (two lines). No response trials (triangles) indicated that the participant should not respond to that trial (Knutson et al., 2008).

Analyses focused on responses to gain trials, and low and high reward magnitudes were collapsed to incorporate more trials in order to produce a more stable average response (Huettel & McCarthy, 2001). No response and loss trials were not included in the analysis. The reward anticipation period was the time between cue onset and target onset (e.g., cue + wait period before target). The contrast of interest for reward anticipation compared anticipation of reward (low and high combined) versus anticipation of non-reward trials.

#### Imaging Analysis

The following text is an automated description of the connectivity analysis, as suggested by the developers, directly from the CONN toolbox: All imaging analyses were performed using CONN (Whitfield-Gabrieli & Nieto-Castanon, 2012)(RRID:SCR_009550) release 21.a (Nieto-Castanon, & Whitfield-Gabrieli, n.d.) and SPM (Penny, Friston, Ashburner, Kiebel, & Nichols, n.d.) (RRID:SCR_007037) release 12.7771.

Preprocessing: Functional and anatomical data were preprocessed using a flexible preprocessing pipeline (Nieto-Castanon, 2020b), including realignment with correction of susceptibility distortion interactions, slice timing correction, outlier detection, direct segmentation and MNI-space normalization, and smoothing. Functional data were realigned using SPM realign & unwarp procedure (Andersson et al., 2001), where all scans were coregistered to a reference image (first scan of the first session) using a least squares approach and a 6 parameter (rigid body) transformation (Friston, Ashburner, Frith, Poline, Heather, & Frackowiak, 1995) and resampled using b-spline interpolation to correct for motion and magnetic susceptibility interactions. Temporal misalignment between different slices of the functional data (acquired in interleaved Siemens order) was corrected following SPM slice-timing correction (STC) procedure (Henson et al., 1999; Sladky et al., 2011) using sinc temporal interpolation to resample each slice BOLD timeseries to a common mid-acquisition time.

Potential outlier scans were identified using ART (Whitfield-Gabrieli, S., Nieto-Castanon, A., & Ghosh, S., 2011) as acquisitions with framewise displacement above 0.5 mm or global BOLD signal changes above three standard deviations (Nieto-Castanon, 2022; Power et al., 2014), and a reference BOLD image was computed for each subject by averaging all scans excluding outliers. Functional and anatomical data were normalized into standard MNI space, segmented into grey matter, white matter, and CSF tissue classes, and resampled to 2 mm isotropic voxels following a direct normalization procedure (Calhoun et al., 2017) using SPM unified segmentation and normalization algorithm (Ashburner, 2007; Ashburner & Friston, 2005) with the default IXI-549 tissue probability map template. Last, functional data were smoothed using spatial convolution with a Gaussian kernel of 6 mm full width half maximum (FWHM).

Denoising: Functional data were denoised using a standard denoising pipeline (Nieto-Castanon, 2020a) including the regression of potential confounding effects characterized by white matter timeseries (5 CompCor noise components), CSF timeseries (5 CompCor noise components), motion parameters and their first-order derivatives (12 factors) (Friston et al., 1996), outlier scans (below 28 factors) (Power et al., 2014), and linear trends (2 factors) within each functional run, followed by bandpass frequency filtering of the BOLD timeseries (Hallquist et al., 2013) between 0.008 Hz and 0.09 Hz. CompCor (Behzadi, Y. et al., 2007; Chai et al., 2012). Noise components within white matter and CSF were estimated by computing the average BOLD signal and the largest principal components orthogonal to the BOLD average, motion parameters, and outlier scans within each subject’s eroded segmentation masks (Behzadi, Y. et al., 2007). From the number of noise terms included in this denoising strategy, the effective degrees of freedom of the BOLD signal after denoising were estimated to range from 68.2 to 80 (average 77.5) across all subjects (Nieto-Castanon, 2022).

#### A priori seed regions

Based on the hypotheses, a seed-to-voxel analysis was performed, using seeds from the left and right nucleus accumbens, putamen, caudate nucleus, and insula. The insula was included as an *a priori* seed region due to its significant group x time interaction with the caudate in the resting-state analysis of this dataset (Hynd et al., 2025). The Automated Anatomical Labelling (AAL) atlas, the default atlas in CONN Toolbox, was used to define the subcortical seeds. Insula seeds—spherical ROIs measuring 10mm—were developed in accordance with Toffoletto et al.’s (2014) review of female brain areas influenced by estrogen (Toffoletto et al., 2014).

#### Planned Analysis

Age was a covariate in all imaging analyses. Reaction time to the target was analyzed using a multilevel model with fMRI visit, group, and cue type as fixed effects, an interaction term for fMRI visit × group, and a random intercept.

Pre-E2 analysis of group differences in functional connectivity during reward anticipation between PO-MDD and control groups utilized a GLM via a seed-to-voxel gPPI procedure in the CONN toolbox. All 8 ROIs mentioned previously were used as seed regions, estimating individual GLMs with voxel-level connectivity measures as dependent variables (one measure per subject) and group as the independent variable. Voxel-level hypotheses were evaluated with multivariate parametric statistics, incorporating random effects across subjects and sample covariance estimation. Results were thresholded using a cluster-forming p < 0.001 voxel-level threshold and a familywise corrected p-FWE < 0.01 cluster-size threshold (Chumbley et al., 2010). Additionally, group (PO-MDD, control) × Time (pre-E2, post-E2) interactions were assessed with another GLM, applying the same seed-to-voxel gPPI procedure and significance thresholds. Significant interactions were further analyzed using post hoc t-tests to explore both between- and within-group connectivity differences.

The relationships between changes in functional connectivity during reward anticipation and PO-MDD symptom reduction over E2 administration were analyzed using a multilevel model including only the PO-MDD group. Self-reported symptoms (at four timepoints) were the dependent variable, and the change in functional connectivity (post-E2 minus pre-E2) served as the moderator of interest. In these analyses, the IDAS Dysphoria subscale and the MASQ-AD were utilized as symptom indicators, aligning closely with the study’s focus on aberrant reward processing. Other fixed effects included visit number, pre-E2 connectivity, the interaction of visit number and pre-E2 connectivity, and the interaction of visit number and change in connectivity.

A random effect variable (random intercept) was included in the model to capture unexplained variance. The Johnson-Neyman technique (Long & Long, 2019) was used on each significant interaction to determine the range in which E2 had a significant influence on symptom change.

## RESULTS

### Clinical Characteristics

Walsh et al. (2025) reported demographic and clinical data for a sample largely overlapping with the current one. There were no significant age differences between groups. The PO-MDD group showed significantly greater reductions in MASQ-AD and IDAS Dysphoria scores from pre-E2 to post-E2 compared to controls (*p*’s < 0.001).

### Target Reaction Times

There were no significant between-group differences in reaction times at pre-E2 or post-E2 for any of the cue types. There was no main effect of group, cue type, and no significant interaction between fMRI visit and group (all *p* > 0.05). However, there was a significant main effect of fMRI visit (t_(3564)_ = −2.41, *p* = 0.02), indicating that reaction times decreased significantly for both groups between the first and second fMRI sessions (see Figure 3).

**Figure 3.**
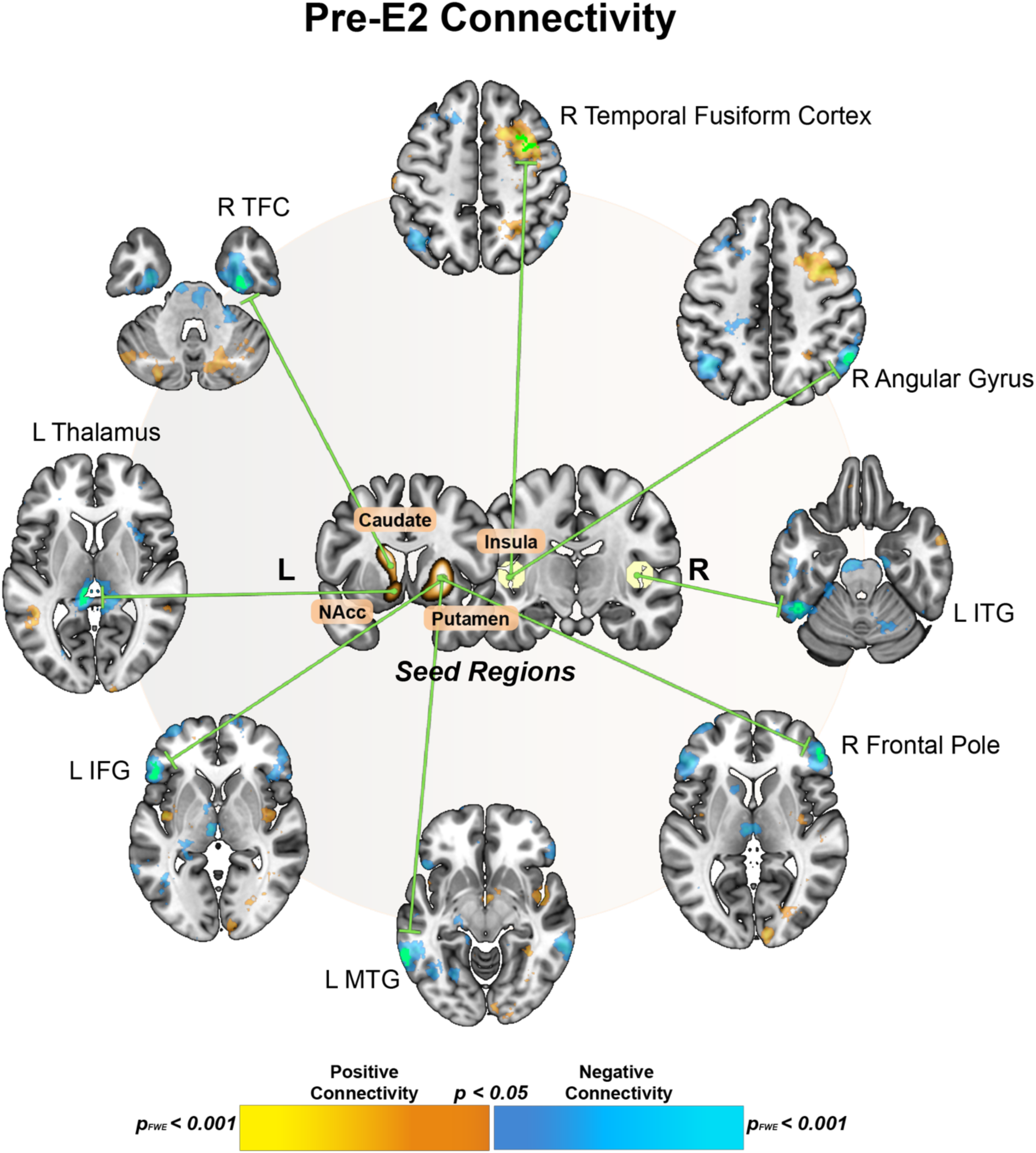
Pre-E2 Connectivity Differences Between PO-MDD and Control Groups. Significant seed regions are depicted in the center of the figure, with green lines illustrating the significant target regions each seed region connects to. Subcortical spheres (caudate nucleus, putamen, and nucleus accumbens) are shown using the AAL atlas regions, while cortical seeds (insula) are depicted with 10mm spherical ROIs. Green target regions passed the pFWE < 0.001 threshold, while other colored regions were significant at the p < 0.05 level, illustrated using the “highlight don’t hide” method (Taylor et al., 2023). L: left. R: right. NAcc: nucleus accumbens. ITG: inferior temporal gyrus. MTG: middle temporal gyrus. IFG: inferior frontal gyrus. TFC: temporal fusiform cortex.

### Aim 1: Pre-E2 Differences

Pre-E2 differences in functional connectivity during reward anticipation were observed in the PO-MDD group compared with the control group across multiple seed regions (see Figure 4). The PO-MDD group exhibited decreased connectivity between the left caudate nucleus and the right temporal fusiform cortex (pFWE < 0.001, k=77). The right putamen showed decreased connectivity with both the left inferior frontal gyrus (pFWE < 0.001, k=88) and the left middle temporal gyrus (pFWE < 0.001, k=75), as well as with the right frontal pole (pFWE = 0.002, k=56). The left nucleus accumbens demonstrated decreased connectivity with the left thalamus (pFWE = 0.001, k=61). In contrast, the left insula displayed increased connectivity with the right middle frontal gyrus (pFWE < 0.001, k=149) but decreased connectivity with the right angular gyrus (pFWE = 0.001, k=61). Lastly, the right insula exhibited decreased connectivity with the left inferior temporal gyrus (pFWE <0.001, k=108).

**Figure 4.**
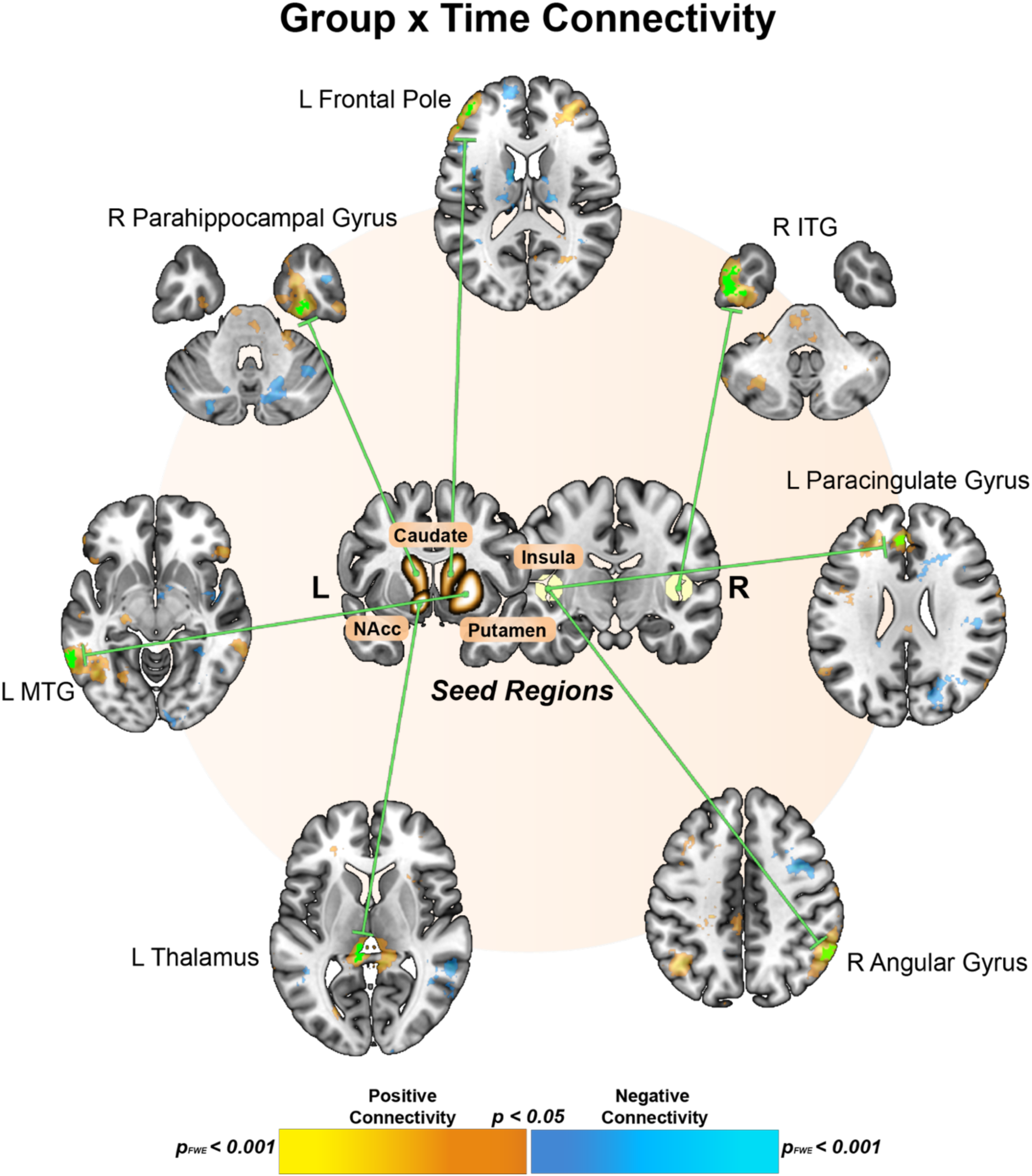
Group x Time Connectivity Interactions. Significant seed regions are depicted in the center of the figure, with green lines illustrating the significant target regions each seed region connects to. Subcortical spheres (caudate nucleus, putamen, and nucleus accumbens) are shown using the AAL atlas regions, while cortical seeds (insula) are depicted with 10mm spherical ROIs. Green target regions passed the pFWE < 0.001 threshold, while other colored regions were significant at the p < 0.05 level, illustrated using the “highlight don’t hide” method (Taylor et al., 2023). L: left. R: right. NAcc: nucleus accumbens. ITG: inferior temporal gyrus. MTG: middle temporal gyrus.

**Figure 5.**
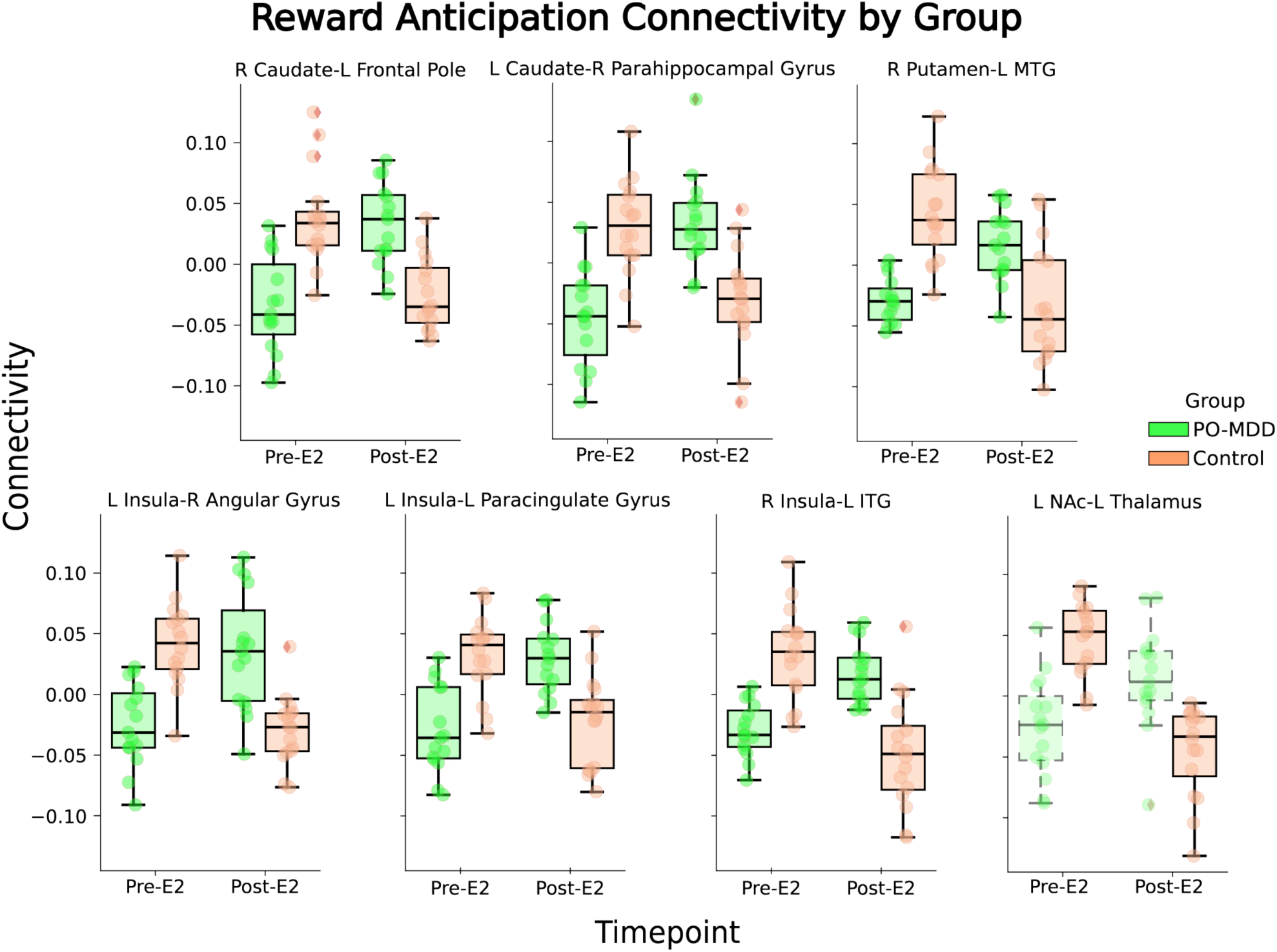
Group × time interactions in reward anticipation connectivity. Boxplots display functional connectivity values across time points (Pre-E2, Post-E2) for each significant interaction, with PO-MDD (green) and Control (orange) groups. All within- and between-group t-tests are significant at p<0.05 except for the PO-MDD within-group (transparent and dashed) t-test for the left nucleus accumbens—left thalamus. R: right. L: left. ITG: inferior temporal gyrus. NAc: nucleus accumbens.

### Aim 2: Group × Time Interactions

Significant Group × Time interactions in functional connectivity were observed across multiple seed regions. In the PO-MDD group, compared with the control group, the right caudate exhibited increased connectivity with the left frontal pole (*p*_FWE_ < 0.001, k=53) and with the right temporal fusiform cortex (*p*_FWE_ < 0.001, k=72). The right putamen demonstrated increased connectivity over time with the left middle temporal gyrus, temporooccipital part (*p*_FWE_ = 0.002, k=56).

The left amygdala demonstrated increased connectivity with the left thalamus (*p*_FWE_ < 0.001, k=77). Likewise, the left nucleus accumbens exhibited increased connectivity with the left thalamus (*p*_FWE_ = 0.001, k=58). The left insula displayed increased connectivity with the right angular gyrus (*p*_FWE_ < 0.001, k=84) and the left/right paracingulate gyrus (*p*_FWE_ < 0.001, k=72). The right insula exhibited increased connectivity with the left inferior temporal gyrus (*p*_FWE_ < 0.001, k=165).

All between- and within-group post hoc t-tests were significant at *p* < 0.05, except for the PO-MDD within-group t-test of left nucleus accumbens–left thalamus connectivity (*p* > 0.05). For all ROI–target pairs, the PO-MDD group showed increased reward anticipation connectivity over time compared with the control group, which showed the opposite effect. See Figures 6 and 7 for a depiction of the results.

**Figure 6.**
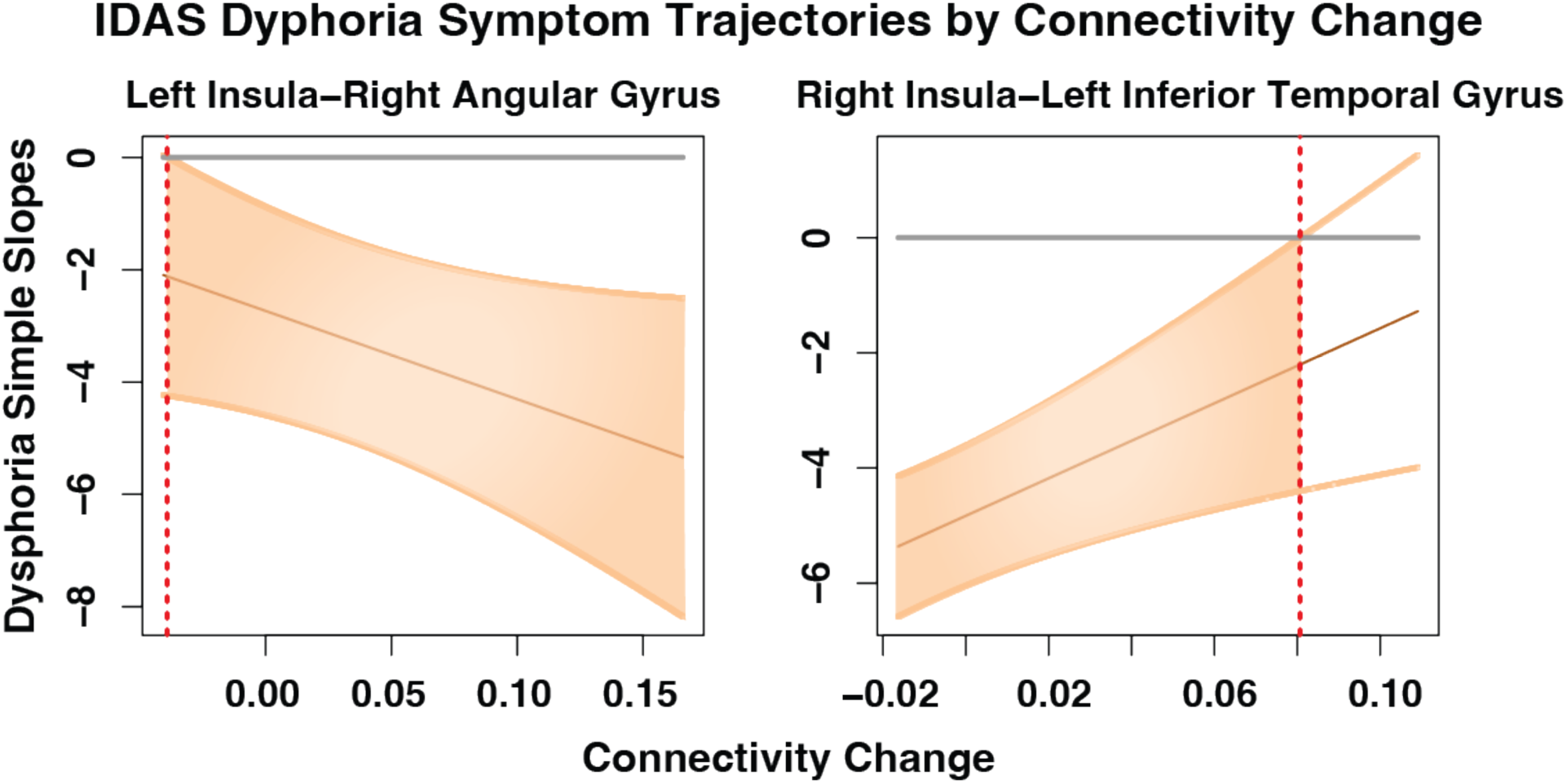
Simple slopes of IDAS Dysphoria symptom trajectory × connectivity change over time in the PO-MDD group. The horizontal gray line represents a slope of 0, indicating no change in symptoms over time. The vertical dotted red line indicates the point at which the interaction effect is no longer significant. Orange-shaded regions indicate statistical significance. For an example interpretation, an increase in the right insula—left inferior temporal gyrus connectivity over time of 0.02 corresponds to a slope of approximately −4 for IDAS Dysphoria symptoms, indicating a steady decrease across E2 administration.

### Aim 3: Associations between Connectivity Changes and Symptom Trajectories

Results from the multilevel model indicated that changes in functional connectivity significantly predicted the trajectory of self-reported dysphoria symptoms over E2 administration. Specifically, connectivity between the left insula–right angular gyrus and the right insula–left inferior temporal gyrus emerged as significant moderators of changes in dysphoria. Reductions in connectivity with E2 administration between the left insula and right angular gyrus were linked to reductions in dysphoria. In contrast, increased connectivity between the right insula and left inferior temporal gyrus was associated with a reduction in dysphoria. Johnson-Neyman analyses further delineated the specific ranges of connectivity change within which these effects were significant, as shown in Figure 6.

## DISCUSSION

This pharmaco-fMRI study explored the impact of transdermal E2 administration on task-based functional connectivity in brain regions related to reward processing among perimenopausal women with PO-MDD and those without PO-MDD (i.e., controls). Our results suggest potential neural mechanisms through which transdermal E2 influences reward processing in PO-MDD.

### Aim 1: Pre-E2 Differences

The current results indicate differences in pre-E2 functional connectivity between the PO-MDD and control groups during the anticipation of reward. It was hypothesized that the PO-MDD group would exhibit lower reward-related connectivity than the control group. During reward anticipation, the hypothesis was supported by the finding that the PO-MDD group exhibited decreased connectivity between the left caudate nucleus and the right temporal fusiform cortex, as well as between the right putamen and multiple cortical regions, including the left inferior frontal gyrus, left middle temporal gyrus, and right frontal pole. The caudate nucleus and putamen are central components of the striatum, which plays a key role in reward anticipation, motivational salience, and habit formation, though these regions also contribute to other cognitive and motor processes (Haruno & Kawato, 2006). Reduced connectivity between these regions and frontal and temporal cortices may indicate impairments in goal-directed behavior and reward expectancy in PO-MDD, consistent with findings from functional connectivity studies of reward dysfunction in major depressive disorder (Admon et al., 2015; Hanneke Geugies et al., 2022; E. Walsh et al., 2017).

Similarly, decreased connectivity between the left nucleus accumbens and the left thalamus further supports the hypothesis of impaired reward-related signaling in PO-MDD. The nucleus accumbens responds strongly to reward consumption and predictability (Berns et al., 2001; Pizzagalli et al., 2009), while the thalamus is involved in relaying sensory and cognitive information to higher-order cortical areas, which is disrupted in MDD and may affect reward-related signaling (Brown et al., 2017). Disrupted connectivity between these regions may reflect deficits in processing reward-relevant cues, which could contribute to anhedonic symptoms. However, it is essential to note that the control group appears to have driven this effect, as the post hoc within-group t-test was not significant for the PO-MDD group, whereas it was significant for the control group.

In contrast to the widespread decreased connectivity shown by the PO-MDD group compared with the control group, the left insula showed increased connectivity with the right middle frontal gyrus but decreased connectivity with the right angular gyrus. In the context of reward processing, the insula is involved in interoceptive awareness and emotional regulation (Menon & Uddin, 2010), while the middle frontal gyrus is implicated in cognitive control as part of the central executive network (Zhu et al., 2022). Increased connectivity between these regions in PO-MDD may reflect compensatory mechanisms to regulate affective responses to anticipated rewards. Reduced connectivity with the angular gyrus, a part of the default mode network, is associated with mapping task-related information (Grabner et al., 2011), which may indicate disruptions in the integration of contextual reward information.

### Aim 2: Group × Time Interactions

It was hypothesized that E2 administration would increase connectivity during reward anticipation in the PO-MDD group between the striatum, insula, and other regions implicated in reward processing impairments in MDD relative to changes observed in the control group. Overall, supporting this hypothesis, the PO-MDD group showed increased connectivity across most seed regions during reward anticipation, while the control group displayed the opposite trend, following E2 administration. Exogenous E2 administration may lead to enhanced reward-related connectivity in PO-MDD, reinforcing the idea that E2 boosts engagement in reward circuits over time.

During reward anticipation, the PO-MDD group exhibited increased connectivity across multiple striatal, limbic, and cortical circuits following E2 administration. In contrast, the control group showed a decrease in connectivity following E2 administration. Specifically, the right and left caudate in the PO-MDD group demonstrated increased connectivity with the left frontal pole and right temporal fusiform cortex, respectively. In the context of reward processing, the caudate is central to reward prediction and motivational salience. In the context of reward processing, the caudate is central to reward prediction and motivational salience. Increased caudate–cortex connectivity following E2 administration may reflect partial normalization of previously observed impairments in PO-MDD, suggesting compensatory recruitment of cognitive control mechanisms during reward anticipation (Amin et al., 2006; Hwang et al., 2015).

The right putamen in the PO-MDD group showed increased connectivity with the left middle temporal gyrus. The left nucleus accumbens also showed increased connectivity with the left thalamus. These regions are critical for decision-making, emotional processing, and reward encoding, among other cognitive and motor functions (Haruno & Kawato, 2006; Zhang et al., 2018). Strengthened connectivity here may indicate adaptive changes in how individuals with PO-MDD process reward cues over time, potentially driven by E2.

Increased connectivity of the left insula with the right angular gyrus and left paracingulate gyrus in PO-MDD following E2 administration may reflect altered integration of cognitive and affective processes during reward anticipation. In the context of reward processing, the insula, involved in interoception, the angular gyrus, serving as a hub for information integration, and the paracingulate gyrus, which contributes to self-referential thinking and reward valuation, may indicate improved reward processing with E2 treatment in hormonally sensitive depression (Paus et al., 1996; Seghier, 2013). Notably, these changes were not observed in the control group, suggesting that the effect is not attributable to E2 alone but instead reflects a targeted antidepressant response on PO-MDD. Furthermore, increased connectivity between the right insula and the left inferior temporal gyrus, another region involved in multisensory integration, may suggest a greater recruitment of sensory-association networks following E2. These paralimbic and cortical regions have also been implicated in prior studies of reward-related connectivity in MDD (E. Walsh et al., 2017) and in the same sample during resting-state (Hynd et al., 2025).

### Aim 3: Connectivity Change Interaction with Symptom Trajectory

The findings from Aim 3 show that alterations in functional connectivity during reward anticipation significantly predict symptom trajectories throughout the administration period. These results align with our hypothesis that notable changes in reward-related connectivity would predict symptom trends during E2 administration. Both increases and decreases in connectivity were predicted to lead to greater symptom improvement over three weeks, depending on the seed—target pair.

During reward anticipation, increased connectivity between the left insula and right angular gyrus was associated with greater symptom improvement. The angular gyrus is implicated in self-referential processing (among other cognitive functions), and its coupling with the insula may reflect more adaptive rumination, although prior work on this topic comes mainly from resting-state studies (Cauda et al., 2011; Kandilarova et al., 2018).

In contrast, decreased connectivity between the right insula and left inferior temporal gyrus was associated with greater symptom improvement. Unlike the other findings, this suggests that decoupling this specific temporal-insular circuit may be adaptive in the context of E2 administration. Decreased connectivity between these regions may facilitate better regulation of affective responses to anticipated rewards, contributing to symptom reduction (Dupont, 2002; Menon & Uddin, 2010).

### Limitations

Limitations of this prospective cohort open-label clinical trial include the modest sample size and the absence of a placebo control, which limits causal inference about E2’s effects. Because the study lacked a placebo or active comparator group, it remains unclear whether the observed effects are unique to E2 or generalize to other treatments for PO-MDD. Future studies should employ a randomized placebo-controlled design, preferably including both a patient and a control group. Additionally, a larger sample size is necessary to validate the findings. Finally, the comparative effectiveness of E2 versus other potential antidepressant treatments, including selective serotonin reuptake inhibitors (SSRI), serotonin-norepinephrine reuptake inhibitors (SNRI), and non-invasive brain stimulation, cannot be evaluated based on the findings of the current study. Future studies are needed to guide treatment decision-making for women presenting with new-onset depression during the menopause transition and the neural mechanisms of action associated with each.

## CONCLUSION

Taken together, these findings suggest 1) PO-MDD is characterized by disruptions in reward-related connectivity, with decreased frontostriatal and limbic connectivity contributing to deficits in reward anticipation, 2) E2 corrects for these deficits by increasing reward-related connectivity in PO-MDD but not controls, and 3) changes in connectivity within paralimbic and prefrontal circuits following E2 administration are associated with symptom reduction PO-MDD, providing a potential neural mechanism of the antidepressant effects of E2. Future research should investigate which women benefit most from E2 and over what time course to enable precision medicine for PO-MDD.

## Data Availability

All data produced in the present study are available upon reasonable request to the authors.

## REFERENCES

Admon, R., Nickerson, L. D., Dillon, D. G., Holmes, A. J., Bogdan, R., Kumar, P., Dougherty, D. D., Iosifescu, D. V., Mischoulon, D., Fava, M., & Pizzagalli, D. A. (2015). Dissociable cortico-striatal connectivity abnormalities in major depression in response to monetary gains and penalties. Psychological Medicine, 45(1), 121–131. 10.1017/s0033291714001123

Amin, Z., Epperson, C. N., Constable, R. T., & Canli, T. (2006). Effects of estrogen variation on neural correlates of emotional response inhibition. NeuroImage, 32(1), 457–464. 10.1016/j.neuroimage.2006.03.013

Andersson, J. L. R., Hutton, C., Ashburner, J., Turner, R., & Friston, K. (2001). Modeling Geometric Deformations in EPI Time Series. NeuroImage, 13(5), 903–919. 10.1006/nimg.2001.0746

Andreano, J. M., & Cahill, L. (2009). Sex influences on the neurobiology of learning and memory. Learning & Memory, 16(4), 248–266. 10.1101/lm.918309

Ashburner, J. (2007). A fast diffeomorphic image registration algorithm. NeuroImage, 38(1), 95–113. 10.1016/j.neuroimage.2007.07.007

Ashburner, J., & Friston, K. J. (2005). Unified segmentation. NeuroImage, 26(3), 839–851. 10.1016/j.neuroimage.2005.02.018

Bayer HealthCare Pharmaceuticals. (2007). Prescribing information Climara (Estradiol Transdermal System).

Behzadi, Y., Restom, K., Liau, J., & Liu, T. T. (2007). A component based noise correction method (CompCor) for BOLD and perfusion based fMRI. Neuroimage, 37(1), 90–101.

Berns, G. S., McClure, S. M., Pagnoni, G., & Montague, P. R. (2001). Predictability Modulates Human Brain Response to Reward. Journal of Neuroscience, 21(8), 2793–2798. 10.1523/JNEUROSCI.21-08-02793.2001

Bore, M. C., Liu, X., Gan, X., Wang, L., Xu, T., Ferraro, S., Li, L., Zhou, B., Zhang, J., Vatansever, D., Biswal, B., Klugah-Brown, B., & Becker, B. (2023). Distinct neurofunctional alterations during motivational and hedonic processing of natural and monetary rewards in depression – a neuroimaging meta-analysis. Psychological Medicine, 1–13. 10.1017/S0033291723003410

Brinton, R. D., Yao, J., Yin, F., Mack, W. J., & Cadenas, E. (2015). Perimenopause as a neurological transition state. Nature Reviews. Endocrinology, 11(7), 393–405. 10.1038/nrendo.2015.82

Bromberger, J. T., & Epperson, C. N. (2018). Depression During and After the Perimenopause: Impact of Hormones, Genetics, and Environmental Determinants of Disease. Obstetrics and Gynecology Clinics of North America, 45(4), 663–678. 10.1016/j.ogc.2018.07.007

Brown, E. C., Clark, D. L., Hassel, S., MacQueen, G., & Ramasubbu, R. (2017). Thalamocortical connectivity in major depressive disorder. Journal of Affective Disorders, 217, 125–131. 10.1016/j.jad.2017.04.004

Calhoun, V. D., Wager, T. D., Krishnan, A., Keri S. Rosch, Rosch, K. S., Seymour, K. E., Nebel, M. B., Mostofsky, S. H., Nyalakanai, P., & Kiehl, K. A. (2017). The impact of T1 versus EPI spatial normalization templates for fMRI data analyses. Human Brain Mapping, 38(11), 5331–5342. 10.1002/hbm.23737

Cao, B., Zhu, J., Zuckerman, H., Rosenblat, J. D., Brietzke, E., Pan, Z., Subramanieapillai, M., Park, C., Lee, Y., & McIntyre, R. S. (2019). Pharmacological interventions targeting anhedonia in patients with major depressive disorder: A systematic review. Progress in Neuro-Psychopharmacology and Biological Psychiatry, 92, 109–117. 10.1016/j.pnpbp.2019.01.002

Cauda, F., D’Agata, F., Sacco, K., Duca, S., Geminiani, G., & Vercelli, A. (2011). Functional connectivity of the insula in the resting brain. NeuroImage, 55(1), 8–23. 10.1016/j.neuroimage.2010.11.049

Chai, X. J., Castañón, A. N., Ongür, D., & Whitfield-Gabrieli, S. (2012). Anticorrelations in resting state networks without global signal regression. NeuroImage, 59(2), 1420–1428. 10.1016/j.neuroimage.2011.08.048

Chavez, C., Hollaus, M., Scarr, E., Pavey, G., Gogos, A., & van den Buuse, M. (2010). The effect of estrogen on dopamine and serotonin receptor and transporter levels in the brain: An autoradiography study. Brain Research, 1321, 51–59. 10.1016/j.brainres.2009.12.093

Chumbley, J., Worsley, K., Flandin, G., & Friston, K. (2010). Topological FDR for neuroimaging. NeuroImage, 49(4), 3057–3064. 10.1016/j.neuroimage.2009.10.090

Deecher, D., Andree, T. H., Sloan, D., & Schechter, L. E. (2008). From menarche to menopause: Exploring the underlying biology of depression in women experiencing hormonal changes. Psychoneuroendocrinology, 33(1), 3–17. 10.1016/j.psyneuen.2007.10.006

Dibonaventura, M. D., Wagner, J.-S., Alvir, J., & Whiteley, J. (2012). Depression, quality of life, work productivity, resource use, and costs among women experiencing menopause and hot flashes: A cross-sectional study. The Primary Care Companion for CNS Disorders, 14(6), PCC.12m01410. 10.4088/PCC.12m01410

Dreher, J.-C., Schmidt, P. J., Kohn, P., Furman, D., Rubinow, D., & Berman, K. F. (2007). Menstrual cycle phase modulates reward-related neural function in women. Proceedings of the National Academy of Sciences, 104(7), 2465–2470. 10.1073/pnas.0605569104

Dupont, S. (2002). Investigating temporal pole function by functional imaging. Epileptic Disorders, 4(S1), S17–S22. 10.1684/j.1950-6945.2002.tb00513.x

First, M. B., & Gibbon, M. (2004). The Structured Clinical Interview for DSM-IV Axis I Disorders (SCID-I) and the Structured Clinical Interview for DSM-IV Axis II Disorders (SCID-II). In Comprehensive handbook of psychological assessment, Vol. 2: Personality assessment (pp. 134–143). John Wiley & Sons, Inc.

Friston, K. J., Ashburner, J., Frith, C. D., Poline, J. B., Heather, J. D., & Frackowiak, R. S. (1995). Spatial registration and normalization of images. Human Brain Mapping. https://onlinelibrary.wiley.com/doi/abs/10.1002/hbm.460030303?casa_token=Rf9UAPGBJ28AAAAA:2umwMw3lL8El30evjsnxT5Ea83JDaEHBsbj5JVCbQIFlEhHZ0jcZ4_84HdYdaDaGSnQuSnX8w8lpChj0

Friston, K. J., Williams, S., Howard, R., Frackowiak, R. S., & Turner, R. (1996). Movement-related effects in fMRI time-series. Magnetic Resonance in Medicine, 35(3), 346–355. 10.1002/mrm.1910350312

Gordon, J. L., Eisenlohr-Moul, T. A., Rubinow, D. R., Schrubbe, L., & Girdler, S. S. (2016). Naturally Occurring Changes in Estradiol Concentrations in the Menopause Transition Predict Morning Cortisol and Negative Mood in Perimenopausal Depression. Clinical Psychological Science, 4(5), 919–935. 10.1177/2167702616647924

Gordon, J. L., & Sander, B. (2021). The role of estradiol fluctuation in the pathophysiology of perimenopausal depression: A hypothesis paper. Psychoneuroendocrinology, 133, 105418. 10.1016/j.psyneuen.2021.105418

Grabner, R. H., Ansari, D., Koschutnig, K., Reishofer, G., & Ebner, F. (2011). The function of the left angular gyrus in mental arithmetic: Evidence from the associative confusion effect.

Hallquist, M. N., Hwang, K., & Luna, B. (2013). The nuisance of nuisance regression: Spectral misspecification in a common approach to resting-state fMRI preprocessing reintroduces noise and obscures functional connectivity. NeuroImage, 82, 208–225. 10.1016/j.neuroimage.2013.05.116

Hanneke Geugies, Nynke A. Groenewold, Maaike Meurs, Bennard Doornbos, Jessica de Klerk-Sluis, Philip van Eijndhoven, Annelieke M. Roest, & Henricus G. Ruhé. (2022).

Decreased Reward Circuit Connectivity During Reward Anticipation in Major Depression. NeuroImage: Clinical, 103226–103226. 10.1016/j.nicl.2022.103226

Haruno, M., & Kawato, M. (2006). Different Neural Correlates of Reward Expectation and Reward Expectation Error in the Putamen and Caudate Nucleus During Stimulus-Action-Reward Association Learning. Journal of Neurophysiology, 95(2), 948–959. 10.1152/jn.00382.2005

Henson, R., Büchel, C., Josephs, O., & Friston, K. (1999). The slice-timing problem in event-related fMRI.

Hidalgo-Lopez, E., Mueller, K., Harris, T. A., Aichhorn, M., Sacher, J., & Pletzer, B. (2020). Human menstrual cycle variation in subcortical functional brain connectivity: A multimodal analysis approach. Brain Structure & Function, 225(2), 591–605. 10.1007/s00429-019-02019-z

Huettel, S. A., & McCarthy, G. (2001). The effects of single-trial averaging upon the spatial extent of fMRI activation. NeuroReport, 12(11), 2411.

Hwang, J. W., Egorova, N., Yang, X. Q., Zhang, W. Y., Chen, J., Yang, X. Y., Hu, L. J., Sun, S., Tu, Y., & Kong, J. (2015). Subthreshold depression is associated with impaired resting-state functional connectivity of the cognitive control network. Translational Psychiatry, 5(11), e683. 10.1038/tp.2015.174

Hynd, M., Gibson, K., Walsh, M., Phillips, R., Prim, J., Eisenlohr-Moul, T., Walsh, E., Dichter, G., & Schiller, C. (2025). Estradiol modulates resting-state connectivity in perimenopausal depression. Journal of Affective Disorders, 371, 253–260. 10.1016/j.jad.2024.11.068

Kandilarova, S., Stoyanov, D., Kostianev, S., & Specht, K. (2018). Altered Resting State Effective Connectivity of Anterior Insula in Depression. Frontiers in Psychiatry, 9, 83. 10.3389/fpsyt.2018.00083

Kenna, H. A., Rasgon, N. L., Geist, C., Small, G., & Silverman, D. (2009). Thalamo-Basal Ganglia connectivity in postmenopausal women receiving estrogen therapy. Neurochemical Research, 34(2), 234–237. 10.1007/s11064-008-9756-z

Knutson, B., Bhanji, J. P., Cooney, R. E., Atlas, L. Y., & Gotlib, I. H. (2008). Neural responses to monetary incentives in major depression. Biological Psychiatry, 63(7), 686–692. Research Support, N.I.H., Extramural Research Support, Non-U.S. Gov’t

Knutson, B., Fong, G. W., Adams, C. M., Varner, J. L., & Hommer, D. (2001). Dissociation of reward anticipation and outcome with event-related fMRI. Neuroreport, 12(17), 3683– 3687. Research Support, U.S. Gov’t, P.H.S.

Long, J. A., & Long, M. J. A. (2019). Package “interactions.” https://interactions.jacob-long.com

Menon, V., & Uddin, L. Q. (2010). Saliency, switching, attention and control: A network model of insula function. Brain Structure & Function, 214(5–6), 655–667. 10.1007/s00429-010-0262-0

Nagy, G. A., Cernasov, P., Pisoni, A., Walsh, E., Dichter, G. S., & Smoski, M. J. (2020). Reward Network Modulation as a Mechanism of Change in Behavioral Activation. Behavior Modification, 44(2), 186–213. 10.1177/0145445518805682

Nieto-Castanon, A. (2020a). FMRI denoising pipeline. In Handbook of functional connectivity Magnetic Resonance Imaging methods in CONN (pp. 17–25). Hilbert Press.

Nieto-Castanon, A. (2020b). FMRI minimal preprocessing pipeline. In Handbook of functional connectivity Magnetic Resonance Imaging methods in CONN (pp. 3–16). Hilbert Press.

Nieto-Castanon, A. (2022). Preparing fMRI Data for Statistical Analysis. 10.48550/ARXIV.2210.13564

Nieto-Castanon, A. & Whitfield-Gabrieli, S. (n.d.). CONN functional connectivity toolbox: RRID SCR_009550, release 21. Retrieved July 20, 2023, from https://www.hilbertpress.org/link-nieto-castanon2021

Pan, P. M., Sato, J. R., Paillère Martinot, M.-L., Martinot, J.-L., Artiges, E., Penttilä, J., Grimmer, Y., van Noort, B. M., Becker, A., Banaschewski, T., Bokde, A. L. W., Desrivières, S., Flor, H., Garavan, H., Ittermann, B., Nees, F., Papadopoulos Orfanos, D., Poustka, L., Fröhner, J. H.,…IMAGEN Consortium. (2022). Longitudinal Trajectory of the Link Between Ventral Striatum and Depression in Adolescence. The American Journal of Psychiatry, 179(7), 470–481. 10.1176/appi.ajp.20081180

Paus, T., Tomaiuolo, F., Otaky, N., MacDonald, D., Petrides, M., Atlas, J., Morris, R., & Evans, A. C. (1996). Human Cingulate and Paracingulate Sulci: Pattern, Variability, Asymmetry, and Probabilistic Map. Cerebral Cortex, 6(2), 207–214. 10.1093/cercor/6.2.207

Penny, W. D., Friston, K. J., Ashburner, J. T., Kiebel, S. J., & Nichols, T. E. (n.d.). Statistical Parametric Mapping: The Analysis of Functional Brain Images—1st Edition. Retrieved July 20, 2023, from https://shop.elsevier.com/books/statistical-parametric-mapping-the-analysis-of-functional-brain-images/penny/978-0-12-372560-8

Pizzagalli, D. A. (2022). Toward a Better Understanding of the Mechanisms and Pathophysiology of Anhedonia: Are We Ready for Translation? American Journal of Psychiatry, 179(7), 458–469. 10.1176/appi.ajp.20220423

Pizzagalli, D. A., Holmes, A. J., Dillon, D. G., Goetz, E. L., Birk, J. L., Bogdan, R., Dougherty, D. D., Iosifescu, D. V., Rauch, S. L., & Fava, M. (2009). Reduced caudate and nucleus accumbens response to rewards in unmedicated individuals with major depressive disorder. The American Journal of Psychiatry, 166(6), 702–710. 10.1176/appi.ajp.2008.08081201

Power, J. D., Mitra, A., Laumann, T. O., Snyder, A. Z., Schlaggar, B. L., & Petersen, S. E. (2014). Methods to detect, characterize, and remove motion artifact in resting state fMRI. NeuroImage, 84, 320–341. 10.1016/j.neuroimage.2013.08.048

Pritschet, L., Santander, T., Taylor, C. M., Layher, E., Yu, S., Miller, M. B., Grafton, S. T., & Jacobs, E. G. (2020). Functional reorganization of brain networks across the human menstrual cycle. NeuroImage, 220, 117091. 10.1016/j.neuroimage.2020.117091

Schiller, C. E., Johnson, S. L., Abate, A. C., Schmidt, P., Rubinow, D. R., David R. Rubinow, & Rubinow, D. R. (2016). Reproductive Steroid Regulation of Mood and Behavior. Comprehensive Physiology, 6(3), 1135–1160. 10.1002/cphy.c150014

Seghier, M. L. (2013). The Angular Gyrus. The Neuroscientist, 19(1), 43–61. 10.1177/1073858412440596

Siobán D. Harlow, Margery Gass, Janet E. Hall, Roger Lobo, Pauline Maki, Robert W. Rebar, Sherry Sherman, Patrick M. Sluss, & Tobie J. de Villiers. (2012). Executive Summary of the Stages of Reproductive Aging Workshop + 10: Addressing the Unfinished Agenda of Staging Reproductive Aging. The Journal of Clinical Endocrinology and Metabolism, 97(4), 1159–1168. 10.1210/jc.2011-3362

Sladky, R., Friston, K. J., Tröstl, J., Cunnington, R., Moser, E., & Windischberger, C. (2011). Slice-timing effects and their correction in functional MRI. NeuroImage, 58(2), 588–594. 10.1016/j.neuroimage.2011.06.078

Taylor, P. A., Reynolds, R. C., Calhoun, V., Gonzalez-Castillo, J., Handwerker, D. A., Bandettini, P. A., Mejia, A. F., & Chen, G. (2023). Highlight results, don’t hide them: Enhance interpretation, reduce biases and improve reproducibility. NeuroImage, 274, 120138. 10.1016/j.neuroimage.2023.120138

Toffoletto, S., Lanzenberger, R., Gingnell, M., Sundström-Poromaa, I., & Comasco, E. (2014). Emotional and cognitive functional imaging of estrogen and progesterone effects in the female human brain: A systematic review. Psychoneuroendocrinology, 50, 28–52. 10.1016/j.psyneuen.2014.07.025

Walsh, E. C., Prim, J. H., Gibson, K., Hynd, M., Phillips, R. D., Dichter, G. S., Nathan, M. D., Lundegard, L., Schiff, L., Bizzell, J., Belger, A., Rubinow, D. R., & Schiller, C. E. (2025). Effects of estradiol administration on brain activation and anhedonia in perimenopausal women: A pharmaco-fMRI study. Journal of Affective Disorders, 378, 340–349. 10.1016/j.jad.2025.01.033

Walsh, E., Carl, H., Eisenlohr-Moul, T., Minkel, J., Crowther, A., Moore, T., Gibbs, D., Petty, C., Bizzell, J., Smoski, M. J., & Dichter, G. S. (2017). Attenuation of Frontostriatal Connectivity During Reward Processing Predicts Response to Psychotherapy in Major Depressive Disorder. Neuropsychopharmacology, 42(4), 831–843. 10.1038/npp.2016.179

Wariso, B. A., Guerrieri, G. M., Thompson, K., Koziol, D. E., Haq, N., Martinez, P. E., Rubinow, D. R., & Schmidt, P. J. (2017). Depression during the menopause transition: Impact on quality of life, social adjustment, and disability. Archives of Women’s Mental Health, 20(2), 273–282. 10.1007/s00737-016-0701-x

Watson, D., & Clark, L. A. (2012). Mood and Anxiety Symptom Questionnaire [Dataset]. 10.1037/t13679-000

Watson, D., O’Hara, M. W., Simms, L. J., Kotov, R., Chmielewski, M., McDade-Montez, E. A., Gamez, W., & Stuart, S. (2007). Development and validation of the Inventory of Depression and Anxiety Symptoms (IDAS). Psychological Assessment, 19(3), 253–268. 10.1037/1040-3590.19.3.253

Whitfield-Gabrieli, S., & Nieto-Castanon, A. (2012). Conn: A functional connectivity toolbox for correlated and anticorrelated brain networks. Brain Connectivity, 2(3), 125–141. 10.1089/brain.2012.0073

Whitfield-Gabrieli, S., Nieto-Castanon, A., & Ghosh, S. (2011). Artifact detection tools (ART) (Version Release Version, 7(19), 11) [Computer software]. https://www.nitrc.org/projects/artifact_detect/

Whitton, A. E., Kumar, P., Treadway, M. T., Rutherford, A. V., Ironside, M. L., Foti, D., Fitzmaurice, G., Du, F., & Pizzagalli, D. A. (2023). Distinct profiles of anhedonia and reward processing and their prospective associations with quality of life among individuals with mood disorders. Molecular Psychiatry, 28(12), 5272–5281. 10.1038/s41380-023-02165-1

Whitton, A. E., Treadway, M. T., & Pizzagalli, D. A. (2015). Reward processing dysfunction in major depression, bipolar disorder and schizophrenia. Current Opinion in Psychiatry, 28(1), 7–12. 10.1097/YCO.0000000000000122

Zhang, F., Peng, W., Sweeney, J. A., Jia, Z., & Gong, Q. (2018). Brain structure alterations in depression: Psychoradiological evidence. CNS Neuroscience & Therapeutics, 24(11), 994–1003. 10.1111/cns.12835

Zhu, Z., Wang, Y., Lau, W. K. W., Wei, X., Liu, Y., Huang, R., & Zhang, R. (2022). Hyperconnectivity between the posterior cingulate and middle frontal and temporal gyrus in depression: Based on functional connectivity meta-analyses. Brain Imaging and Behavior, 16(4), 1538–1551. 10.1007/s11682-022-00628-7

