## Supplement for "Estradiol’s Effect on Reward-Related Functional Connectivity in Perimenopause-Onset Major Depressive Disorder"

*Supplementary Table 1.*

| ROI Seed | Target (x, y, z) | Brodmann Area | Target Name | Target Cluster Size | GLM p-FDR |
|---|---|---|---|---|---|
| Nucleus Accumbens | | | | | |
| *Left* | -08, -36, +08 | | Left Thalamus | 58 | < 0.01** |
| Putamen | | | | | |
| *Right* | -62, -52, -06 | 37 | Left Fusiform | 56 | < 0.01** |
| Caudate Nucleus | | | | | |
| *Left* | +38, -18, -34 | 36 | Right Parahippocampal Gyrus | 72 | < 0.001*** |
| | +54, +06, +28 | 6 | Right Pre-motor cortex/ supplementary motor area | 37 | < 0.05* |
| *Right* | -52, +36, +20 | 46 | Left Frontal Pole | 53 | < 0.01** |
| Insula | | | | | |
| *Right* | -50, -06, -38 | 20 | Left Inferior Temporal Gyrus | 165 | < 0.001*** |

Reward anticipation group x time target seed description table.

The following analyses include additional data and results that were not part of the primary aims of the manuscript; however, they still provide relevant contributions to the literature. This includes additional seed regions in the reward anticipation analysis and examination of the reward receipt phase of the MID task.

Aim 1. Baseline Differences

Reward Receipt

In the PO-MDD group, the ACC exhibited decreased connectivity with the right putamen ($p_{FWE} = 0.003$, k=50) when compared to the control group. Similarly, the L-DLPFC in the PO-MDD group showed decreased connectivity with the precuneus ($p_{FWE} = 0.001$, k=58), while the R-DLPFC demonstrated a decrease in connectivity with the right precentral gyrus ($p_{FWE} = 0.001$, k=86). Additionally, both the right caudate and left caudate in the PO-MDD group displayed decreased connectivity with the right cerebellum ($p_{FWE} < 0.001$, k=86; $p_{FWE} < 0.001$, k=76) relative to the control group. Finally, the left amygdala in the PO-MDD group exhibited increased connectivity with the left lateral occipital cortex ($p_{FWE} < 0.001$, k=112) compared to the control group. All results survived voxel-wise thresholding at $p < 0.001$ and cluster-level correction at $p_{FWE} < 0.01$. See Supplemental Figure 1.

*Supplemental Figure 1.*

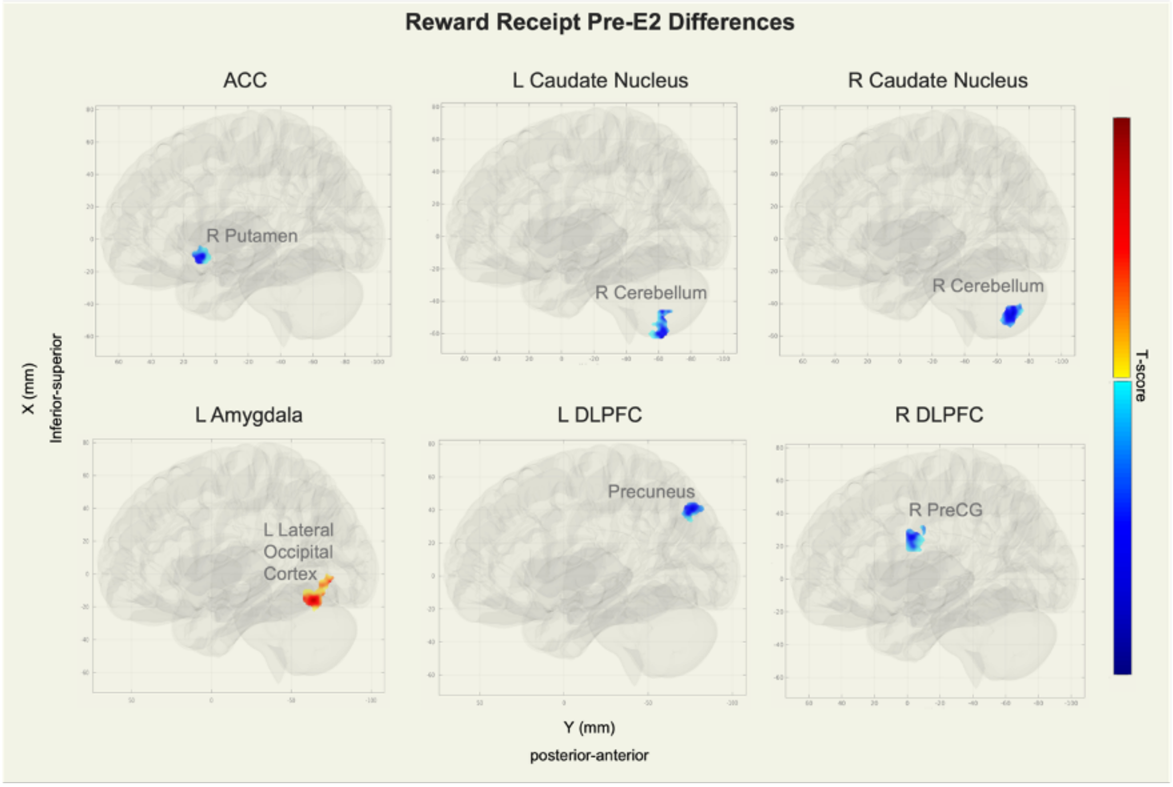


*Clusters showing significant connectivity differences in reward receipt (Pre-E2) between PO-MDD and Control groups (PO-MDD |>| Control). Warmer colors (red) indicate more positive*

*connectivity during reward anticipation in the PO-MDD group, while cooler colors (blue) indicate more negative connectivity in the PO-MDD group compared with the control group. All clusters are significant at the cluster-forming p < 0.001 voxel-level threshold and a familywise corrected p-FWE < 0.01 cluster-size threshold. T-scores range from lightest color to darkest color, 3.60 to 5.95 (red) or -3.60 to -5.95 (blue), respectively. R: right. L: left. PreCG: precentral gyrus.*

Aim 2. Group x Time Interactions

Reward Anticipation

The left amygdala demonstrated increased connectivity with the left thalamus ($p_{FWE}$ < 0.001, k=77). The L-DLPFC (MNI = -9, +54, +30) showed decreased connectivity over time in the PO-MDD group with a posterior portion of the left middle frontal gyrus (pFWE = 0.002, k=56, MNI = -36, +30, +30), referred to hereafter as "left posterior DLPFC" to avoid confusion with its seed region.

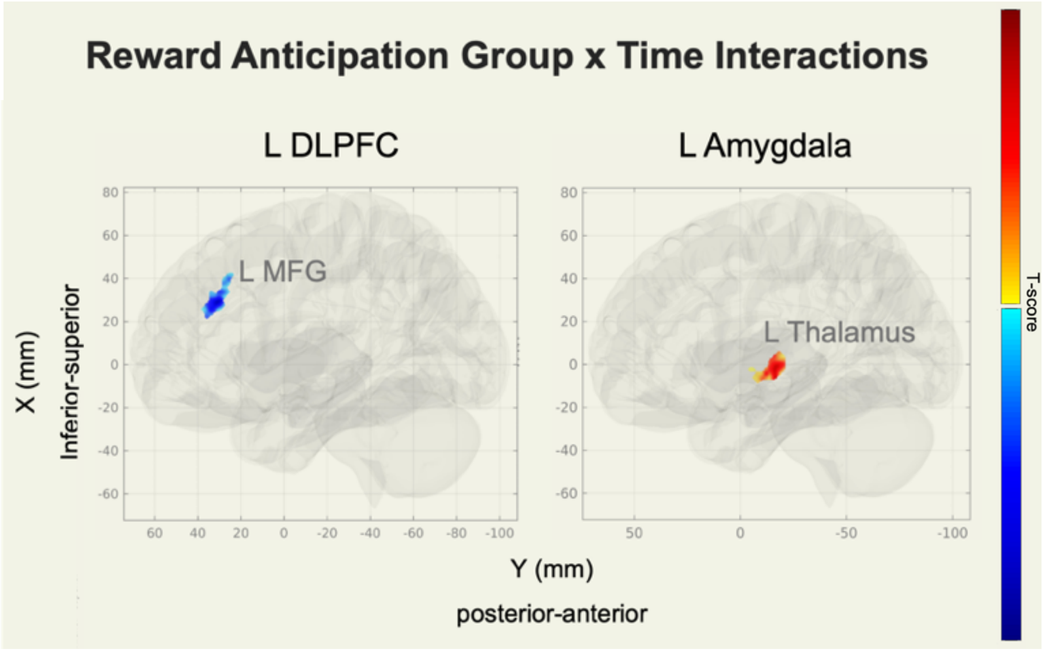


*Supplemental Figure 2.*

*Significant group × time interactions in connectivity during reward anticipation (PO-MDD |>| Control). Warmer colors (red) indicate greater increases in connectivity over time in the PO-MDD group relative to the control group, while cooler colors (blue) indicate greater decreases. All clusters are significant at the cluster-forming p < 0.001 voxel-level threshold and a familywise corrected p-FWE < 0.01 cluster-size threshold. T-scores range from lightest color to* *darkest color, 3.60 to 5.95 (red) or -3.60 to -5.95 (blue), respectively. R: right. L: left. MFG: middle frontal gyrus (left posterior DLPFC).*

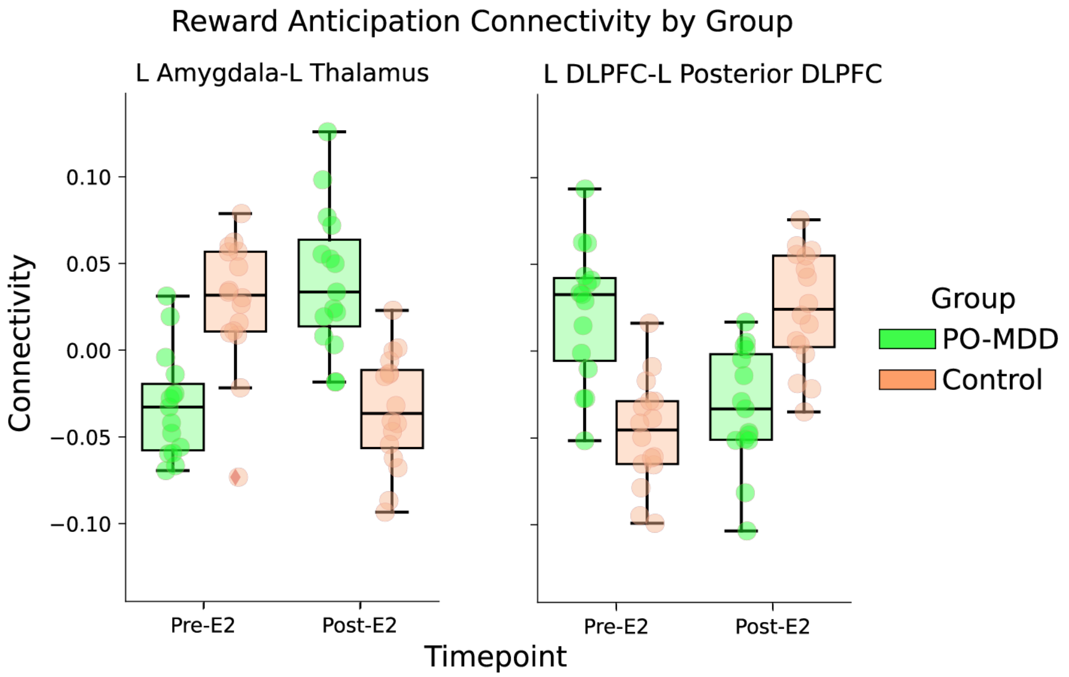


*Supplemental Figure 3.*

*Group × time interactions in reward receipt connectivity. Boxplots display functional connectivity values across time points (Pre-E2, Post-E2) for each significant interaction, with PO-MDD (green) and control (orange) groups. All within- and between-group t-tests are significant at p<0.05. R: right. L: left.*

Reward Receipt

The L-DLPFC demonstrated decreased connectivity with a posterior portion of the left middle frontal gyrus ("left posterior DLPFC"; $p_{FWE} < 0.001$, k=136, MNI coordinates = -30, +28, +28) and increased connectivity to the right postcentral gyrus ($p_{FWE} < 0.001$, k=113) over time. The R-DLPFC showed increased connectivity with the left central opercular cortex/precentral gyrus ($p_{FWE} < 0.001$, k=277) and the right precentral gyrus ($p_{FWE} < 0.001$, k=188). Additionally, the right amygdala exhibited significantly increased connectivity with the right precentral gyrus ($p_{FWE} <0.001$, k=105). All results were significant at a voxel-wise threshold of $p < 0.001$, with a cluster-size threshold corrected at $p_{FWE} < 0.01$. Moreover, all between- and within-group posthoc t-tests were significant at $p < 0.05$. See Supplemental Figures 4 and 5 for a depiction of results.

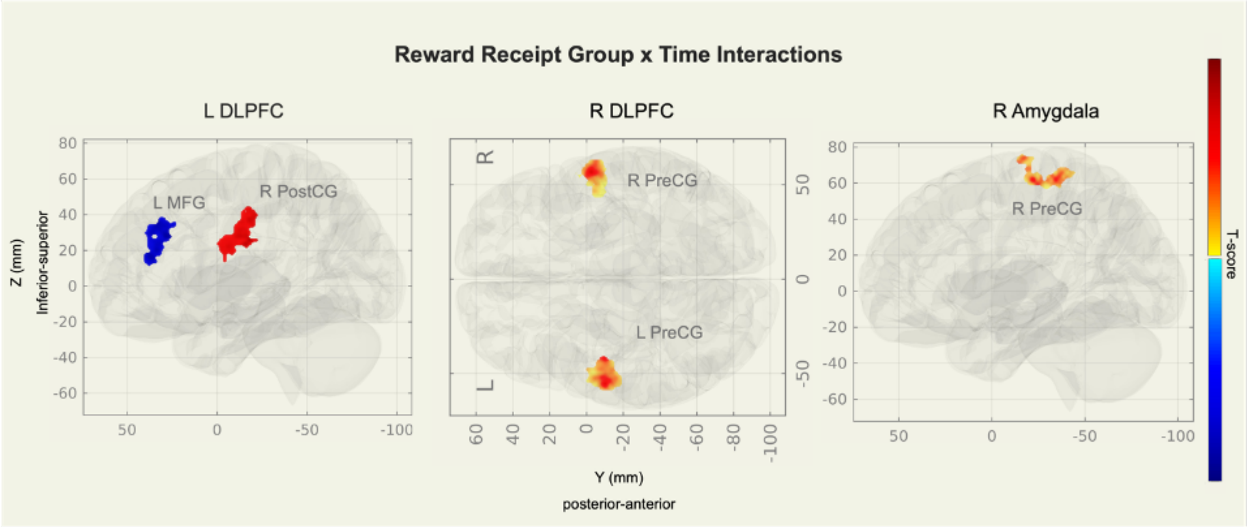


*Supplemental Figure 4.*

*Significant group × time interactions in connectivity during reward receipt (PO-MDD |>| Control). Warmer colors (red) indicate greater increases in connectivity over time in the PO-MDD group relative to the control group, while cooler colors (blue) indicate greater decreases. All clusters are significant at the cluster-forming p < 0.001 voxel-level threshold and a familywise corrected p-FWE < 0.01 cluster-size threshold. T-scores range from lightest color to darkest color, 3.60 to 5.95 (red) or -3.60 to -5.95 (blue), respectively. R: right. L: left. PostCG: postcentral gyrus. MFG: middle frontal gyrus (left posterior DLPFC). PreCG: precentral gyrus.*

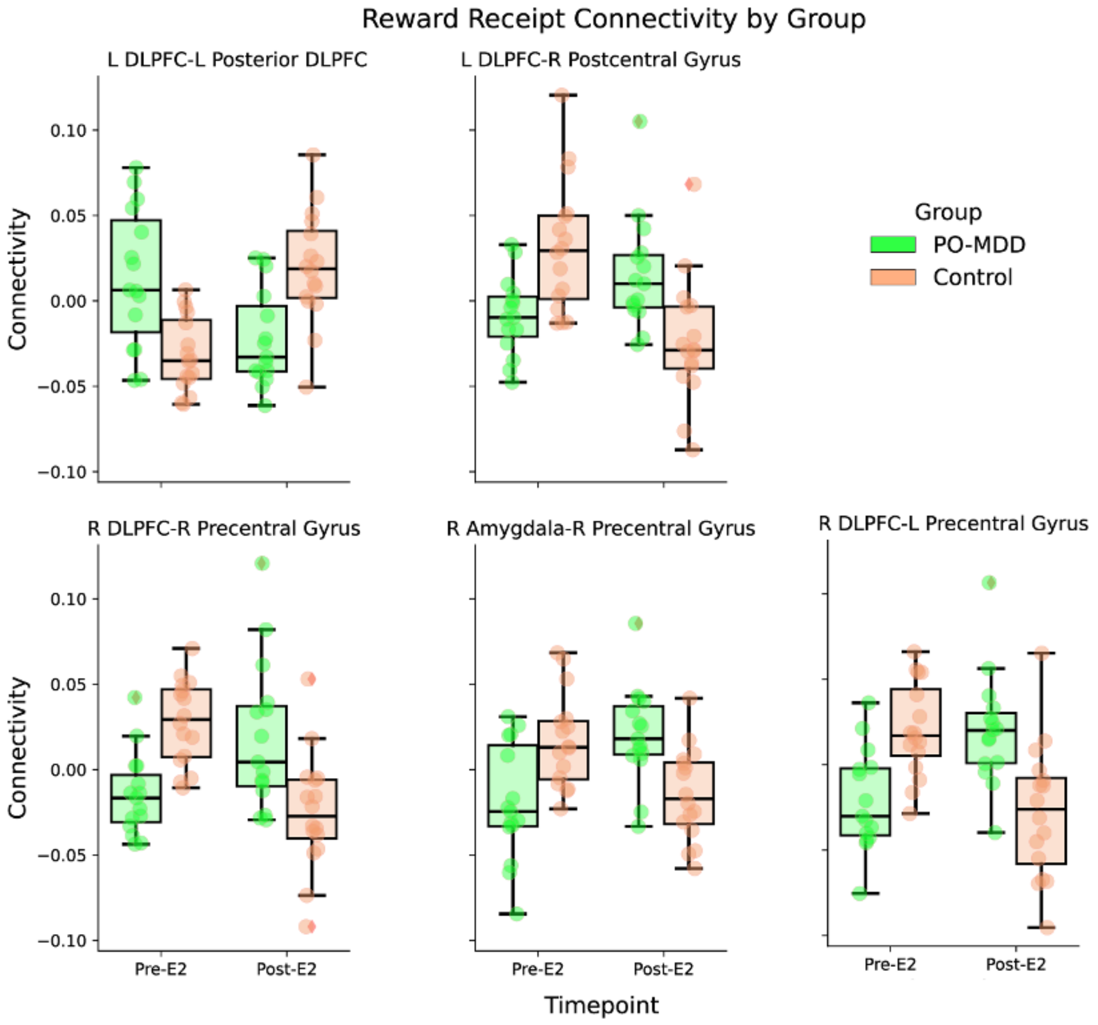


*Supplemental Figure 5.*

*Group × time interactions in reward receipt connectivity. Boxplots display functional connectivity values across time points (Pre-E2, Post-E2) for each significant interaction, with PO-MDD (green) and control (orange) groups. All within- and between-group t-tests are significant at $p<0.05$. R: right. L: left.*

Aim 3: Connectivity Change Interaction with Symptom Trajectory

Reward Anticipation

Results from the multilevel model indicated that changes in functional connectivity during reward anticipation significantly predicted the trajectory of self-reported dysphoria symptoms over administration. Specifically, two additional ROI—target pairs emerged as significant moderators of symptom change. Decreased connectivity between the left amygdala and left thalamus was associated with a steeper slope of symptom reduction over E2 administration, as was decreased connectivity between the L-DLPFC and left posterior DLPFC.

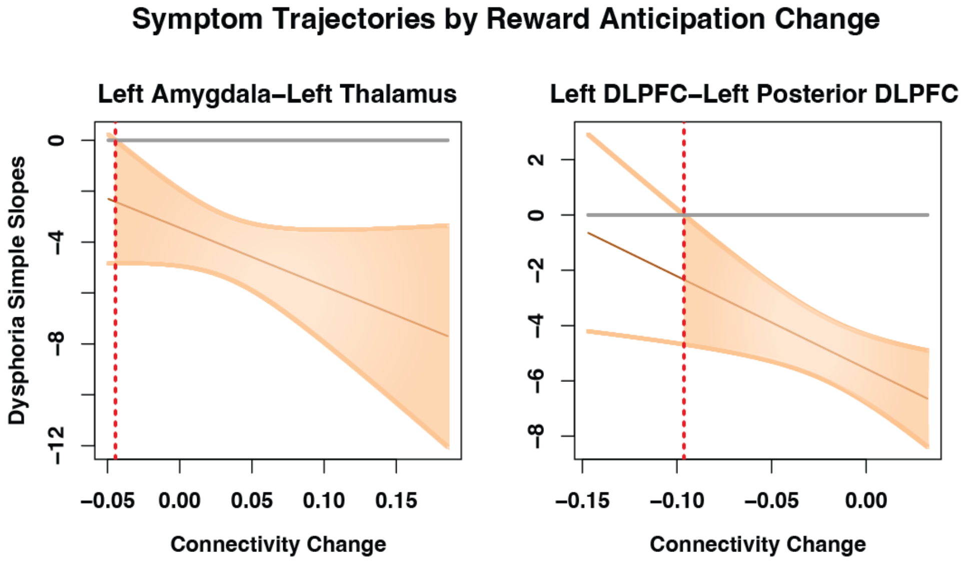


*Supplemental Figure 6.*

*Simple slopes of IDAS Dysphoria symptom trajectory × reward anticipation connectivity change over time in the PO-MDD group. The horizontal gray line represents a slope of 0, indicating no change in symptoms over time. The vertical dotted red line marks the point where the interaction effect is no longer significant. Orange-shaded regions indicate statistical significance. For example, an increase in the left amygdala–left thalamus connectivity over time of 0.10 corresponds to a slope of approximately -6 for IDAS Dysphoria symptoms, indicating a steady decrease across E2 administration.*

Reward Receipt

Decreased connectivity between the left DLPFC and the left posterior DLPFC was associated with more favorable symptom trajectories. Specifically, individuals with minimal or decreased connectivity showed faster improvement in both dysphoria (IDAS) and anhedonia (MASQ-AD) over E2 administration. The Johnson-Neyman intervals indicate that the relationship between connectivity change and symptom reduction was significant at lower levels of connectivity change, as shown in Supplemental Figure 7. No mediation analyses were significant.

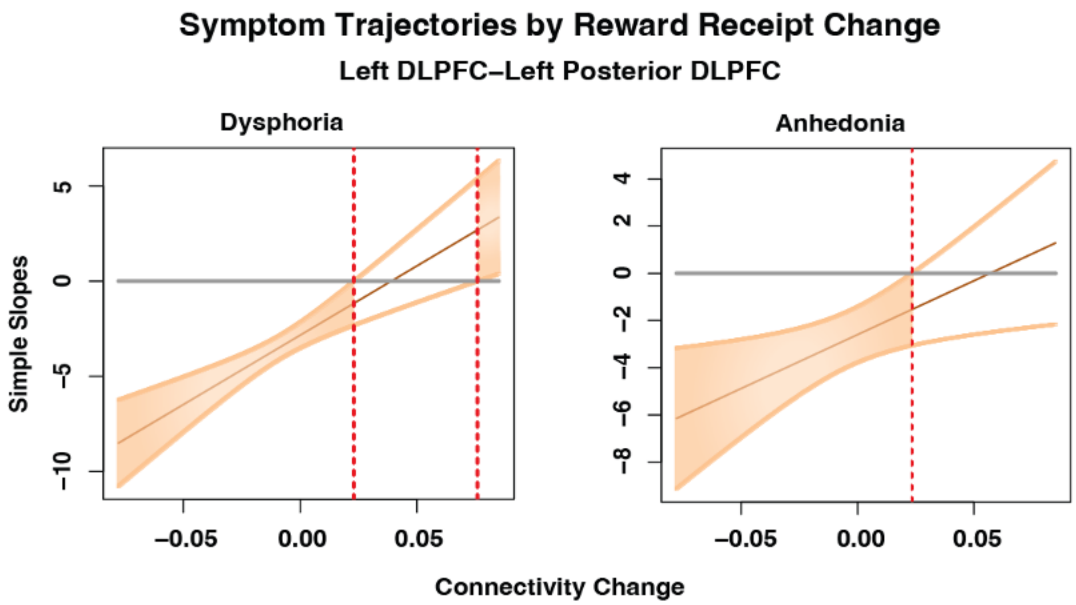


*Supplemental Figure 7.*

*Simple slopes of IDAS Dysphoria and MASQ-AD symptom trajectory × reward receipt connectivity change over time in the PO-MDD group. The horizontal gray line represents a slope of 0 (no symptom change over time). The vertical dotted red line marks the point where the interaction effect is no longer significant. Orange-shaded regions indicate statistical significance. For example, a decrease in left DLPFC—left middle frontal gyrus connectivity over time of 0.05 ("-0.05" in the figure) corresponds to a slope of approximately -7 for IDAS Dysphoria symptoms, indicating a steady decrease across E2 administration.*
